# Planning ahead for research participation: survey of public and professional stakeholders’ views about the acceptability and feasibility of advance research planning

**DOI:** 10.1101/2023.04.25.23289103

**Authors:** Victoria Shepherd, Kerenza Hood, Fiona Wood

## Abstract

**Introduction:** To date, anticipatory planning in the UK has focused on supporting people who anticipate periods of impaired decisional capacity to express their wishes about their future care through processes such as advance care planning. Other countries have extended anticipatory planning to include mechanisms for people to prospectively express their preferences about research participation. Advance research planning (ARP) could extend people’s autonomy and ensure that ‘proxy’ decisions about research are based on their wishes and preferences.

**Objectives:** To explore a range of public and professional stakeholders’ views about the acceptability and feasibility of planning for future research participation and identify barriers and facilitators to implementing ARP.

**Design:** Cross-sectional survey

**Main outcomes:** Acceptability and feasibility of ARP

**Participants:** Between November 2022 and March 2023, two groups of stakeholders (members of the public including people living with capacity-affecting conditions and their carers; researchers and other professionals) were invited to participate in a cross-sectional survey via multiple recruitment routes. Online questionnaires were used to capture the perspectives of the two groups.

**Results:** Responses from members of the public (n=277) and professionals (n=50) were analysed using descriptive statistics and content analysis. Introducing ARP in the UK was supported by 97% of public contributors and 94% of professionals, who recommended it include the person’s general wishes about research, specific types of studies if known, and who should make decisions on their behalf. Challenges include how ARP takes account of changes in individuals’ preferences or circumstances and protecting their rights and interests. Implementation barriers include the potential time, complexity, and cost involved. These may be addressed by embedding ARP in existing anticipatory planning pathways and aligning it with other research enrolment activities. Relationships and trust have a key role, including underpinning who supports the delivery of ARP, how they are trained, and when it is undertaken.

**Conclusions:** There are high levels of support for implementing ARP in the UK. Further research should explore practical barriers and stakeholder concerns and identify any unintended consequences. ARP interventions should be developed alongside training and other resources. Activities should focus on public awareness campaigns, and engaging policymakers and other stakeholders.

**Strengths and limitations of this study:**

1. This is the first exploration of the acceptability and feasibility of advance research planning in the UK
2. Questions were based on previous similar surveys conducted in other countries which ensured prior validation and enables international comparison
3. The study included a wide range of members of the public including people living with conditions that may affect decision-making in the future, and professionals including researchers, research ethics committee members, and healthcare practitioners
4. The non-random selection of participants and inability to track non-responders may have resulted in potential participation bias
5. Participants were predominantly white and had some prior involvement in research, therefore their views may not be representative of more diverse groups or those with less experience of research

## Introduction

More than 920,000 people in the UK are living with dementia, and this is expected to rise to over a million by 2024 (1). Alongside a range of other conditions and disabilities, dementia contributes to the approximately 2 million people in the UK who have significantly impaired decision-making (2). There is an increasing focus on supporting people living with conditions such as dementia and cancer to discuss their preferences about future care and treatment options prior to any loss of capacity. Advance care planning (ACP) is viewed as a way of enabling people with capacity to think about the meanings and consequences of different future scenarios and discuss their goals and preferences with family members and their healthcare providers (3). It encourages them to identify a proxy decision-maker(s) and to record and regularly review any preferences, so that their preferences can be taken into account should they, at some point, be unable to make their own decisions (3). ACP forms part of NICE guidelines for end-of-life care for adults (4), dementia (5), and decision-making and mental capacity (6), and NICE has produced a website of resources on advance care planning for social care (7). However, despite a drive to embed research into care pathways being a key strategic aim in the UK (8), ACP discussions and other formal processes such as Lasting Power of Attorney (LPA) arrangements do not currently extend to preferences and proxy decisions about research participation in the UK.

Conducting research with people who have significantly impaired capacity relies on ‘proxy’ decision-makers to make decisions about participation on their behalf. In the UK, proxy decisions made on behalf of people who lack capacity to consent should be based on what the person’s wishes and feelings about taking part in the study would be (9,10) and should not be contrary to ‘an advance decision … or any other form of statement’ (10). However, people rarely discuss their preferences about research and so it can be difficult for families to make a decision about whether their relative should participate in a research study, and many experience an emotional and decisional burden as a result (11). Previous studies suggest that proxies’ views often differ from what the potential participants would want (12), including a tendency to underestimate the willingness of older adults to participate in research (13). Knowing the person’s wishes may help families or, in the event there is no-one available to act in a personal capacity, a member of their care team, acting as consultees or legal representatives in the event of a loss of capacity (11). It may also contribute to better inclusion in trials of people with impaired capacity to consent as this group is frequently excluded from trials (14,15) – primarily due to the ethical and legal complexities around consent (16).

Advance research planning (ARP) is a broad process which enables people to express and document their preferences about research, and has been proposed as a means to overcome the challenges associated with proxy decision-making for research (17). Other countries have established differing processes for planning ahead for research participation in the event of a loss of capacity. For several decades, legal provisions in the US and in Canada have enabled people to make an advance research directive - a document in which they can specify their wishes about future research participation (18). Since 2007, Australia’s National Statement on Ethical Conduct in Human Research has enabled a formal process for researchers to discuss and document views on future research participation with participants who anticipate periods of cognitive impairment (19). More recently, there have been moves to integrate advance research directives into the European legal framework (20), with legislation awaiting implementation in Switzerland (18). Germany has gone one stage further, with recent changes in legislation that now requires an advance research directive to be in place in order for people lacking capacity to be included in research that does not offer any personal benefit, although this was introduced without any public or expert debate about the implications of these directives (21).

Outside the UK, the feasibility of ARP has been explored in populations including older people (12,22), people living with dementia (21,23), palliative care (24), as well as acute conditions such as stroke (25). ARP has support from organisations such as Alzheimer Europe who encourage the use of advance directives to record peoples’ wishes to participate (or not participate) in research as it respects their right to self-determination and their desire to do something constructive which may eventually benefit others with a similar medical condition (26). It is thought that ARP can promote and extend the autonomy and self-determination of people who wish to plan for future incapacity, help ensure that proxy decision-making reflects the values and wishes of those lacking capacity to consent, and support the inclusion in research of those who have expressed their prior interest in participation (27). It may also reduce the burden experienced by proxies when making a decision on behalf of others (28). However, ARP raises a number of ethical and practical concerns about *what* types of research should ARP be considered for, what information should ARP include and in what format, *when* is the best time for public or patients to undertake ARP and when is the best time to convey that information to others), *who* should be approached to undertake ARP and which groups of professionals should be involved), and *how* should ARP be implemented in practice and which kind of safeguards might need to be in place (22).

In 2009 the Nuffield Council on Bioethics recommended research to explore the feasibility of developing a (non-binding) advance statement on research participation which could influence decisions on research participation after loss of capacity (Recommendation 18) (29). More recently, recommendations for research involving people near the end of life in the UK suggested that clinicians should engage patients in conversations about research participation and document their preferences and wishes, and people who are likely to lose capacity should be asked to designate a consultee to provide an opinion on their participation in a study (30). However, despite the introduction of ARP in other jurisdictions, no studies have explored stakeholders’ views about ARP in the UK. As part of a larger research programme (CONSULT (31)), we conducted the CONSULT-ADVANCE study to explore a range of public and professional stakeholders’ views about the acceptability and feasibility of ARP in the UK and identify the barriers and facilitators to implementing ARP. For the purposes of this study, ARP is broadly viewed as a mechanism to enable people to express their preferences about research rather than to provide ‘advance consent’ for a specific study (22,32) or focusing solely on the creation of an advance research directive, although it could potentially include these activities.

## Methods

### Design, setting and participants

This cross-sectional study used an online survey to explore attitudes towards ARP from a broad range of public and professional stakeholders. The public stakeholder group includes people who have personal experience of either living with dementia or another condition which may affect their ability to consent to research, are a family member or friend of someone with such a condition, or a member of the public interested in research or advance planning. The professional stakeholder group includes researchers or other professionals with an interest in research into capacity-affecting conditions or who have an interest in advance planning. Participants were not eligible if they were unable to understand English sufficiently to comprehend the study information and complete the survey.

Ethical approval was obtained from the School of Medicine Research Ethics Committee at Cardiff University (SMREC ref. 22.84) prior to commencing the survey.

### Procedure

There were different recruitment pathways for the two types of stakeholder groups. Details about the study were shared with public stakeholders via social media platforms (e.g Twitter) and community or support groups (e.g Parkinson’s UK, Stroke Association). They were also recruited through Join Dementia Research which is an online registry that enables volunteers with memory problems or dementia, carers of those with memory problems or dementia and healthy volunteers to sign up and register their interest in taking part in research. Researchers and other professional stakeholders with an interest in dementia or another condition which may affect their capacity were invited to participate via social media platforms (e.g Twitter) and research networks (e.g MRC-NIHR

Trials Methodology Research Partnership, British Society for Gerontology). Research funders’ databases of current and previously funded studies (e.g NIHR portfolio) were also searched to identify researchers who are working in relevant areas, and other professional stakeholders such as research ethics committee members were invited through organisations such as the Health Research Authority.

### Questionnaires

Two questionnaires were developed based on previous surveys conducted with public and professional stakeholders in other countries (e.g (17,23)) and adapted for the UK context and the aims of this study. The questionnaires were designed to capture the different, although corresponding, perspectives of the two groups. Data collection was via an online survey tool (Qualtrics) between November 2022 and March 2023. Participants were provided with a link to access the online questionnaire that was relevant to the stakeholder group they identified with (public or professional). Each contained a home page with participant information about the study, followed by the questionnaire pages. Participants were asked to tick a box at the start of the survey to confirm that they consented to participate. The professionals’ survey consisted of two sections containing questions about their demographics and seeking their views about advance research planning (Appendix 1). The survey for members of the public had an additional section seeking their views about research participation (Appendix 2). Both versions had a combination of multiple-choice questions, Likert-type scales, and free-text responses and the option of providing contact details if they were willing to be contacted about taking part in an interview for the next stage of the study.

### Patient and public involvement

The survey questions and information pages were reviewed by a lay advisory group who support the wider research programme which this study forms a part. They also reviewed the findings and contributed to their interpretation and the implications.

### Data analysis

Descriptive analysis was used to report the demographic characteristics and responses. A supplementary file provides frequency distributions for all response options. Content analysis was performed on free-text responses (33). Responses were coded using qualitative data analysis software (NVivo) and common themes were identified and reported thematically.

## Results

### Demographic characteristics of participants

A total of 277 participants responded to the survey for members of the public. Public stakeholder characteristics are shown in **Table 1**. A total of 50 professionals responded to the survey for researchers and other professionals. Professional stakeholder characteristics are shown in **Table 2** below.

**Table 1.** Characteristics of public stakeholder survey participants

| Participant characteristic | n=277 (%) |
| --- | --- |
| **Gender** |  |
| Female | 184 (66%) |
| Male | 92 (33%) |
| Other | 1 (0.4%) |
| **Age** |  |
| 18-24 | 7 (3%) |
| 25-34 | 12 (4%) |
| 35-49 | 22 (8%) |
| 50-64 | 96 (35%) |
| 65+ | 140 (51%) |
| **Geographical location** |  |
| England | 191 (69%) |
| Wales | 41 (15%) |
| Scotland | 32 (12%) |
| Northern Ireland | 9 (3%) |
| Other | 4 (1%) |
| **Ethnicity~** |  |
| White | 268 (96%) |
| Asian/Asian British | 5 (2%) |
| Mixed/Multiple ethnic groups | 1 (0.4%) |
| Black/African/Caribbean/Black British | 0 |
| Other ethnic group | 3 (1%) |
| **Stakeholder group** |  |
| Family member/friend of someone with an impairing condition | 87 (31%) |
| Member of the public | 83 (30%) |
| Experience of living with an impairing condition | 67 (24%) |
| Other | 40 (14%) |
| **Previously participated in research** |  |
| Yes | 166 (60%) |
| No | 104 (38%) |
| Other | 7 (3%) |
| **Previously participated in advance planning activities** |  |
| Yes | 161 (58%) |
| No | 111 (40%) |
| Other | 5 (2%) |
| **Area of interest^** |  |
| Dementia | 236 (85%) |
| Parkinson's disease | 53 (19%) |
| Stroke | 34 (12%) |
| Palliative or end of life care | 26 (9%) |
| Huntington's disease | 3 (1%) |
| Other | 17 (6%) |

~Questions and response categories were based on ONS guidance for collecting data about ethnic group
^Participants could select more than one option

**Table 2.** Characteristics of professional stakeholder survey participants

| **Participant characteristic** | **n=50 (%)** |
| --- | --- |
| **Length of time involved in research (years)** |  |
| 0-5 | 14 (28%) |
| 6-10 | 9 (18%) |
| 11+ | 26 (52%) |
| Other | 1 (2%) |
| **Geographical location** |  |
| England | 18 (36%) |
| Wales | 28 (56%) |
| Scotland | 0 |
| Northern Ireland | 0 |
| Other | 4 (8%) |
| **Ethnicity~** |  |
| White | 48 (96%) |
| Asian/Asian British | 1 (2%) |
| Mixed/Multiple ethnic groups | 0 |
| Black/African/Caribbean/Black British | 1 (2%) |
| Other ethnic group | 0 |
| **Stakeholder group** |  |
| Researcher with an interest in capacity-affecting condition(s) | 23 (46%) |
| HCP* caring for people with capacity-affecting conditions | 8 (16%) |
| Researcher with an interest in advance planning | 6 (12%) |
| Research ethics committee member | 2 (4%) |
| Researcher with an interest in ethics and law | 1 (2%) |
| Other | 10 (20%) |
| **Role includes approaching participants about research** |  |
| Yes | 34 (68%) |
| No | 15 (30%) |
| Other | 1 (2%) |
| **Role includes advance planning activities** |  |
| Yes | 15 (30%) |
| No | 32 (64%) |
| Other | 3 (6%) |
| **Area of interest^** |  |
| Dementia | 24 (48%) |
| Parkinson's disease | 2 (4%) |
| Stroke | 7 (14%) |
| Palliative or end of life care | 16 (32%) |
| Huntington's disease | 3 (6%) |
| Other | 11 (22%) |

~Questions and response categories were based on ONS guidance for collecting data about ethnic group
\* HCP = health care professional
^Participants could select more than one option

## Public stakeholders’ views about research participation

### Willingness to participate in research in the event of loss of capacity

Public stakeholder participants were asked to imagine that they had dementia or another condition affecting their memory or understanding and had been identified as being able to take part in a research study but at that point in time they are unable to make their own decision about taking part in the study. Participants were provided with brief descriptions of different types of research studies and asked how much they agree or disagree with statements about their willingness to participate on a 5-point Likert-type scale from strongly agree to strongly disagree.

Participants showed a high degree of willingness to participate in research should capacity be lost, with a mean of 90% of participants stating they either strongly agree or agree they would be willing to participate, although this varied by study type (see **Table 3)** with lower levels of agreement for those involving an experimental medicine (65%) compared with those involving tests of memory or thinking (96%) or body measurements (96%).

**Table 3.** Public stakeholders’ willingness to participate in research by study type

| I would be willing to be included in a research study that involves<sup>s</sup> | Strongly agree | Agree | Neither disagree nor agree | Disagree | Strongly disagree |
| --- | --- | --- | --- | --- | --- |
| a) Asking me questions in a survey or interview (*e.g asking about my experiences or opinions*) | 178 (64%) | 85 (31%) | 5 (2%) | 8 (3%) | 1 (0.4%) |
| b) Observing my behaviour (*e.g watching how I act if I listen to music*) | 152 (55%) | 104 (38%) | 14 (5%) | 6 (2%) | 1 (0.4%) |
| c) Testing my memory or thinking (*e.g asking me to draw a picture or remember specific words*) | 173 (62%) | 92 (33%) | 7 (3%) | 4 (1%) | 1 (0.4%) |
| d) Giving me psychological therapy (*e.g counselling for anxiety or depression*) | 136 (49%) | 93 (34%) | 31 (11%) | 12 (4%) | 5 (2%) |
| e) Giving me physiotherapy (*e.g moving my arms or legs, massaging my muscles*) | 155 (56%) | 89 (32%) | 24 (9%) | 8 (3%) | 1 (0.4%) |
| f) Giving me experimental medicine (*e.g a drug that might help my condition*) | 75 (27%) | 106 (38%) | 61 (22%) | 22 (8%) | 13 (5%) |
| g) Taking x-rays or scans of my body (*e.g to help researchers see how my condition is affecting my brain*) | 155 (56%) | 97 (35%) | 17 (6%) | 5 (2%) | 3 (1%) |
| h) Taking measurements about my body (*e.g my weight, blood pressure*) | 171 (62%) | 96 (35%) | 6 (2%) | 4 (1%) | 0 (0%) |
| j) Putting something on my body, like a bracelet, that keeps track of information (*e.g how much time I spend in bed*) | 152 (55%) | 98 (35%) | 17 (6%) | 10 (4%) | 0 (0%) |
| k) Taking a sample of my blood or other body fluid for **genetic research** | 162 (58%) | 92 (33%) | 11 (4%) | 7 (3%) | 5 (2%) |
| *(e.g to find out if I and my relatives have a gene that increases the risk of getting dementia)* |  |  |  |  |  |
| l) Taking a sample of my blood or other body fluid for **non-genetic research** *(e.g to find out if my blood shows I had an infection in the past that increases my risk of a condition)* | 164 (59%) | 92 (33%) | 11 (4%) | 6 (2%) | 4 (1%) |
| m) Looking at my personal records, such as medical records or test results held at my GP practice or hospital *(e.g to study how a past illnesses might be related to my condition)* | 167 (60%) | 94 (34%) | 12 (4%) | 1 (0%) | 3 (1%) |
| n) Accessing stored samples of my blood, body fluids or other tissues *(e.g If I had blood taken in the past for another reason, researchers might ask the hospital for access to that blood for study)* | 165 (60%) | 91 (33%) | 13 (5%) | 4 (1%) | 4 (1%) |

§ Participants were asked to imagine a scenario where they had dementia or another condition and could take part in a research study but were not able to make their own decision at that time about taking part in the study. They were asked how much they agreed or disagreed with the statements. Questions adapted from Ries et al 2019 (34)

### Willingness to participate in research with differing benefit profiles

When asked about their willingness to take part in research that may or may not benefit them to varying degrees, 97% (n=266) of public stakeholder participants were willing to take part in research that benefits them directly, 96% (n=265) were willing to take part in research that may not have direct benefit for them but could help others with the same condition, and 90% (n=247) were willing to take part in research that may not benefit them or others with the same condition but could help researchers to understand other diseases or health problems.

## Public and professional stakeholders’ views about advance research planning

### Interest in undertaking advance research planning

Public stakeholder participants were asked how interested they would be in taking part in ARP if it were available. Of the responses from 274 participants, 97% (n=267) were very or somewhat interested, 0.4% (n=1) were either not very or not at all interested, and 2% (n=6) described themselves as being unsure. Those who expressed either uncertainty or disinterest stated that this was either because they preferred someone else to make decisions about their participation in research, that they did not think it was important to express their wishes about taking part in future research, or that they were aware that their views might change over time and therefore views expressed now might not be the same in the future.

### Importance of advance research planning

Professional stakeholder participants were asked how important they thought it was for public/patients to undertake ARP, if it was possible for them to do so. Of the responses, 94% (n=46) thought it was either very important or somewhat important for public/patients to do so, 4% (n=2) thought it was either not very important or not at all important, and 2% (n=1) were unsure. Of those who expressed either uncertainty or thought it not so important, explanations included that they thought another process would better support people to express their wishes for taking part in future research (e.g it should be included in generic advance decision making), or that another process would better support people to express their wishes for taking part in future research (e.g the level of detail required means that advance research planning is unlikely to be successful and individual decisions should be made at the time).

### Acceptability of advance research planning

When asked which types of research ARP was appropriate for, 52% (n=26) of professionals thought all types of research, whilst 46% (n=23) thought it was appropriate for only some types of research with least support for interventional studies involving medicinal products, and 2% (n=1) were unsure. When asked which groups of people ARP might be most appropriate for, professionals were supportive of all groups listed, including people living with conditions such as dementia, Parkinson’s, or Huntington’s disease, and those at high risk of developing these conditions, as well as people at risk of acute medical events such as stroke, and older people in general. They also proposed other groups that they thought ARP would be appropriate for such as people with intellectual disabilities and people with terminal illnesses including children and young people.

### Timing of advance research planning

Participants were asked when ARP should be undertaken. There was most support for it to occur at the same time as other planning processes (e.g when having ACP discussions or making LPA arrangements) with 87% (n=236) of members of the public and 85% (n=39) of professionals either strongly agreeing or agreeing with this. There was also support for ARP to be timed with being approached about a specific study (e.g when joining a research registry, entering an initial observational stage of an interventional study) from 71% (n=193) of public and 72% (n=33) professionals. There was some support for ARP taking place immediately after someone is diagnosed as having (or being at risk of) a condition that might affect their capacity in the future by 66% (n=178) of public and 67% (n=31) professionals, with similar levels of support for ARP occurring at opportunistic or ad hoc times (e.g at any point when motivated or interested in doing so - as with decisions about organ donation) by 66% (n=180) of public and 63% (n=29) professionals. Most participants thought it should be reviewed at different timepoints (e.g at regular timepoints or when there are changes in their personal circumstances or clinical condition) with 91% (n=246) of members of public and 89% (40) of professionals either strongly agreeing or agreeing with this.

### Content of advance research planning

ARP might cover different content, and participants were presented with a range of options. Almost all participants either strongly agreed or agreed that it should include nominating who makes decisions on the person’s behalf (e.g naming a particular person such as their spouse/partner) by 95% (n=255) of members of public and 91% (n=42) of professionals, and should include the person’s general wishes about research they would or would not want to participate in (e.g studies related to their condition only or any study they are eligible for) by 94% (n=254) of public and 98% (n=45) of professionals, and the person’s wishes about what specific types of research they would or would not want to participate in (e.g observational or interventional, anticipated benefits and risks involved, involving specific procedures such as blood tests or scans) 96% (n=257) of public and 98% (n=45) of professionals.

### How advance research planning should inform decisions about research

ARP can be used to inform a decision about whether someone should take part in a research study in different ways. Participants were less supportive of whether wishes expressed through ARP should be considered legally binding (i.e they should be followed regardless of what others involved in the process think) (public 46% n=124, professionals 36% n=16), compared with being considered directive (i.e they should directly inform the decision although they do not have to be followed if there are reasonable views otherwise) (public 77% n=206, professionals 84% n=38), with a medium level of support for it being considered advisory (i.e they can help or contribute to the decision) 59% (n=160) of members of public and 69% (n=31).

### What form advance research planning should take

Participants were asked what that form ARP should take. Participants were most supportive of it being a formal process which is documented by the individual and a formal copy shared with others (e.g similar to an advance statement about wishes and care preferences) (public 81% n=217, professionals 89% n=40), compared to it being either an informal discussion which is then written down by the individual themselves and/or by the professional involved in their care records (public 46% n=123, professionals 33% n=15), or a legal process in which wishes about research are documented in a legal document which is then registered (e.g similar to the process for LPA) (public 43% n=116, professionals 29% n=13).

### Who should be involved in advance research planning

Different people could be involved in the process of ARP. Participants thought that someone who is involved in making decisions with or for the person should be involved (e.g a spouse or adult child, someone with Power of Attorney) (public 95% n=255, professionals 80% n=36) and a doctor or other healthcare professional (HCP) who is part of the person’s healthcare team (public 65% n=175, professionals 60% n=27), with less support for a doctor or other research professional who is part of a research team (public 57% n=152, professionals 49% n=23).

## Views about barriers and facilitators to implementing advance research planning

Participants were asked for their views about any barriers to implementing ARP, and what could help support it. Many participants described it as an important area and something they would like to see introduced. Some provided specific examples of where it may have been useful such as during COVID-19 trials, or in emergency trials where a nominated or independent HCP is involved in making a decision about participation. A number of key themes were identified from the free text comments.

### Need for information and understanding

People potentially taking part in ARP, family members, researchers and healthcare and legal professionals need tailored and accessible information about what ARP is and how it might help with decisions about research. Guidance is needed about how to complete documents/forms (e.g advance research directive) and any other arrangements (e.g including in LPA) and how it should be shared and who with. This may need to be accompanied by explanations about research including different types of research, arrangements for decisions about participation, and how decision-making may change over time. Public awareness raising campaigns are also needed, and using case studies or examples might be helpful, supported by public involvement.

### Optimal timing depending on stage of life and illness trajectory

Early ARP was widely supported which would enable people to engage in discussions at a time when they are best able to express their preferences. This may be at a point when someone receives a diagnosis, or as part of ageing-based activities. However, people may be reluctant to do so if it seemed less relevant to them or it could be potentially distressing for people to consider their future loss of capacity, or it may be a time when they are already receiving a lot of information. Dementia-related issues around apathy and agnosia/reluctance to accept a diagnosis may also present challenges. Participants emphasised that individual preferences and cultural influences would need to be considered.

### Practical challenges and proposed solutions

Time, complexity, cost, and accessibility were all suggested as practical barriers to ARP. This might be especially so for family carers who may already be responsible for managing current care requirements and supporting other future planning activities. How ARP is delivered and who by, the training they receive, having adequate funding to support this, and building in flexibility whilst reducing complexity were all suggested as ways of overcoming these barriers. Support may also be needed for people with communication or other disabilities. There may be challenges around the level of specificity possible/required, for example it may be difficult to predict what types of studies may be offered in the future. Whilst health or social care professionals or researchers may not be required to be involved, their support may help people to make a more informed choice.

Researchers and other professionals highlighted issues around being able to access an individual’s advance research plan including where it would be stored or recorded (e.g a registry, somewhere it could be accessed in an emergency, ‘version control’). Researchers and other professionals were also concerned it may become a ‘tick box’ exercise at the expense of personalised conversations, or that it might become a requirement. There is also a need to ensure equity by not excluding those who do not have a family member or friend they could nominate to make decisions on their behalf.

### Integrating advance research planning into existing pathways and processes

Participants highlighted the need to minimise the burden of ARP for all those involved, especially as it might not be seen as a priority. They suggested that integrating ARP into other future planning activities might help reduce the burden and improve uptake. Embedding conversations about ARP into existing care pathways was also viewed as helpful, although the challenges of introducing this into an already pressured healthcare system was identified. Discussions about future research preferences could also be incorporated into recruitment processes for existing studies or when signing up to research registries.

### Relationships and the importance of trust

Participants were concerned that the wishes and preferences expressed through ARP might not always be understood or respected by family members when making decisions about participation. Having family members engaged in ARP discussions was seen as important by participants. This included practical support to help people access and understand information about the process, as well as ensuring that family members were aware of the existence and contents of those discussions/documents. Trust was seen as a key underpinning of the whole process – including with HCPs and researchers who might be involved.

### Adapting to changing situations and ensuring safeguarding

Participants recognised that people expressing their wishes through ARP may change their mind at a later date, or their circumstances might change. Being able to revisit and review their preferences was seen as important but was seen as potentially challenging due to changes in the person’s cognition over time. Public stakeholders often expressed concerns that preferences stated in advance might mean that someone was involved in research even when it caused them distress. Having processes in place to prevent this and reassuring people that their wellbeing would be prioritised was seen as essential.

## Discussion

This study found high levels of support for introducing ARP in the UK, including 97% of public contributors and 94% of researchers and other professionals. Participants thought that ARP should be a formal process with documents completed by the individual and then shared with others, and preferably with the involvement of a family member. They thought an advance research plan should include indications of the person’s general wishes about research, as well as details of any specific types of research they would or would not want to participate in, and who they would like to make decisions on their behalf. When asked how ARP should be used, participants thought it should directly inform decisions about research participation but should not be legally binding meaning that it would not have to be followed if there were reasonable views otherwise.

Whilst this is the first study to explore the acceptability and feasibility of ARP in the UK, a previous study found that family members with experience of acting as research proxy were supportive of ARP, including the introduction of a process to nominate someone to make future decisions about research and to extending the existing arrangements for a Health and Welfare LPA (35). Our findings reflect studies in other jurisdictions which show widespread support for various forms of ARP (17,21–23). As found in our study, older people in Australia (17) and the US (36) demonstrated a high level of altruism through their willingness to be involved in research during future periods of impaired capacity that may not necessarily directly benefit them but could help others or improve understanding of other health conditions.

The study also identified a number of factors affecting future implementation of ARP in the UK, many of which correspond with those identified in a previous Australian study (37). Barriers to uptake of ARP identified by both groups of stakeholders included the time, complexity, cost that may be involved, and how accessible the process would be. There were concerns from public stakeholders about how ARP might sufficiently allow for changes in individuals’ preferences and their changing circumstances, and that safeguarding processes would be needed to protect individuals’ rights and interests. Factors facilitating implementation included embedding ARP in existing anticipatory planning pathways/activities such as advance care planning, organ donation, and LPA arrangements, and aligning it with other research activities such as joining research registries or when enrolling in existing studies. This aligns with recent conceptualisations of advance care planning as a continuum, along which a range of different preparation and planning activities may take place under the broad ‘umbrella’ of care planning (38). When viewed as a continuum, the optimal timing for ARP was thought to depend on individuals’ circumstances and the process must take account of this, their illness trajectory where relevant, and be tailored to their communication needs. Relationships and trust were seen as fundamental to the aims and process of ARP, including who supports the delivery of ARP. The need for training and resources for all groups involved in ARP was emphasised, along with the importance of activities to raise public awareness, and engagement with policymakers and other stakeholders.

## Strengths and limitations

The study involved a range of public and professional stakeholders (including people living with capacity-affecting conditions and their family members) whose interest included neurodegenerative conditions such as dementia, acute conditions such as stroke, and areas such as palliative and end of life care. Limitations included our recruitment approach which meant that we were unable to track those who did not respond to the invitation and a majority of participants had prior involvement in research, therefore participants’ responses may not be representative of people with less experience of research and those of non-responders. Participants were predominantly white, and so the findings may not reflect the perspectives of more diverse groups or of people who were unable to complete the online survey due to cognitive impairment, additional language or communication needs, or digital exclusion.

## Areas for future research

From our findings and the previous international studies exploring ARP, a number of uncertainties remain. Legislative and research governance frameworks vary between jurisdictions, therefore the process of advance planning for research participation needs to be contextualised in line with the UK legislative and policy environment. There are legal uncertainties about how (or if) it would align with mental capacity legislation across the UK (e.g Mental Capacity Act 2005 in England and Wales (10), Adults with Incapacity (Scotland) Act 2000 (39)) and clinical trials regulation (e.g Medicines for Human Use (Clinical Trials) Regulation (9)). The legal and ethical implications of preferences expressed through ARP activities, including the validity of ‘advance consent’ for specific studies, requires further exploration.

There is the risk that introducing the opportunity to express future wishes about research participation via ARP may lead to this becoming a legal requirement in order to involve people who lack capacity to consent (as is the case for some types of research in Germany (21)) which would disadvantage people who are unable to or do not wish to take part in ARP, which would particularly exclude groups such as people with a learning disability and those least likely to have family members to involve in the process. Further research is needed to explore these and other unintended consequences such as the potential for a breach of rights if individuals are included in research that they would not wish to participate in (for example in emergency research where an ARP was not known or accessible), or where the only option for accessing a treatment is through a clinical trial. Additionally, other policy and legal implications should be explored such as the intersection between ARP and other anticipatory planning arrangements such as LPA, advance decisions to refuse treatment (ADRT) and ‘do not attempt resuscitation’ (DNACPR) decisions. There is also a need to explore the views of diverse communities about the acceptability and feasibility of ARP, particularly with under-served groups who may have differing perspectives on trust and autonomy for example (38).

There is a need to develop interventions to support ARP, such as templates for creating an advance research directive (23,40), which could then be evaluated in future research. Further research is needed to understand stakeholders’ information and resource needs, identify potential delivery routes and the training requirements of those involved, and explore other context-specific implementation factors. The optimal timing, implementation route and associated guidance may need to vary depending on the context - for example this may differ for people diagnosed with a condition such as dementia from those with other life-limiting conditions, or those wishing to engage in more general anticipatory planning. Given that our previous research has identified the low number of studies that include adults with impaired capacity, even in conditions where there may be high levels of cognitive impairment in the target population (14), there are also broader issues about the need to match people expressing an interest in taking part in research and there being a reasonable number of studies that are open for participation. More research is needed to explore whether ARP might help address the challenges of conducting research with adults lacking capacity to consent (16) and support proxies to make better quality participation decisions that are more informed and based on the person’s preferences (41) and so reduce the decisional and emotional burden they experience (11).

## Conclusion

Members of the public, including people living with capacity-affecting conditions, are willing to participate in research if they lack capacity to provide their own consent, even if the study is not intended to benefit them. Members of the public, as well as researchers and other professionals, express high levels of support for implementing ARP in the UK. Ensuring that ARP is embedded in existing advance care planning pathways and research activities is key to overcoming the challenges to its introduction, and that there is appropriate support and training for those involved. The findings from this study can be used to develop ARP interventions and context-specific resources to support people to express their future wishes about research. More research is needed to identify the practical barriers to implementation and to explore the ethico-legal implications with respect to the UK legislative and policy environment. Future activities should also include raising public awareness about ARP and engaging with policymakers and other stakeholders.

## Supporting information

Appendix 1. CONSULT-ADVANCE study questionnaire for professionals

Appendix 2. CONSULT-ADVANCE study questionnaire for members of the public

Supplementary File 1 CONSULT-ADVANCE Study Protocol v1.0 03.11.2022

Supplementary File 2 Additional survey data

CROSS checklist_CONSULT-ADVANCE survey

## Data Availability

The dataset generated and used in this study is available through submission of a data request to the Centre for Trials Research at https://www.cardiff.ac.uk/centre-for-trials-research/about-us/data-requests

## Ethics approval and consent to participate

This study was approved by the Cardiff University School of Medicine Research Ethics Committee (SMREC ref. 22.84). All study participants provided consent prior to participation in the survey.

## Patient consent for publication

Not applicable.

## Authors’ contributions

VS conceived the study, and VS, KH, and FW designed the study. VS recruited study participants and led the analysis. VS, KH and FW interpreted the data. VS drafted the manuscript. All authors critically revised the manuscript and approved the final version.

## Acknowledgements

We would like to thank the organisations who shared the details of the study with their networks (including Join Dementia Research, Stroke Association and Parkinson’s UK), the participants who generously gave their time to participate in this survey, and the lay advisory group who provide invaluable insight and support for this research programme.

