## Appendix 1. CONSULT-ADVANCE study questionnaire for professionals for "Planning ahead for research participation: survey of public and professional stakeholders’ views about the acceptability and feasibility of advance research planning"

### **Part A. About you**

**Which group best describes your main interest in this area?**

- Researcher with an interest in capacity-affecting condition(s)
- Researcher with an interest in advance planning
- Researcher with an interest in ethics and law
- Healthcare professional caring for people with capacity-affecting conditions
- Research ethics committee member
- Other (*please specify*)

**Is your main interest in this area in relation to?**

- Dementia
- Stroke
- Parkinsons disease
- Huntington’s disease
- Palliative care or end of life care
- Other (*please specify*)

**Where do you work?**

- England
- Northern Ireland
- Scotland
- Wales
- Other (*please specify*)

**What is your ethnic group?**

(Choose one option that best describes your ethnic group or background)

- White
- Mixed / Multiple ethnic groups
- Asian / Asian British
- Black / African / Caribbean / Black British
- Other ethnic group

**How long have you been involved in research?**

- 0-5 years
- 6-10 years
- 11+ years
- Other (*please specify*)

**Does your role include approaching participants to take part in research?**

- Yes my role includes approaching participants
- No my role does not include approaching participants
- Other (*please specifiy*)

**Does your role include any advance planning activities e.g supporting people to create an advance directive/statement or set up Power of Attorney, advance care planning)?**

- Yes my role includes advance planning activities
- No my role does not include advance planning activities
- Other (*please specifiy*)

### **Part B. Your views about Advance Research Planning**

**Advance Research Planning** is a process where individuals can express their wishes about being involved in research studies in the future.

They take part in the process of Advance Research Planning at a time when they have capacity to consider their options and make decisions.

It might include expressing **what** their wishes are about being involved in different types of research, and **who** they would like to be involved in making a decision on their behalf.

If they later lose capacity to consent to a study, Advance Research Planning will tell people such as their doctor, a family member/friend, or a researcher what their wishes are.

| **Q1. If it were possible for public/patients to undertake Advance Research Planning, how important do you think it is?** | 1) Very important *(skip next question)* 2) Somewhat important *(skip next question)* 3) Unsure *(skip next question)* 4) Not very important 5) Not at all important |
| --- | --- |
| Why do you not think it is important for people to undertake Advance Research Planning? | 1) I do not think supporting people to express their wishes for taking part in future research is important 2) I do not think processes such as Advance Research Planning will help people to express their wishes for taking part in future research 3) I think another process would better support people to express their wishes for taking part in future research (*please specify*) 4) Other (*please specify*) |

| **Q2. In your view, what types of research might Advance Research Planning be most appropriate for? Tick all that apply** | 1) All types of research 2) Some types of research 3) Not appropriate for any type of research 4) Unsure |
| --- | --- |
| Which types of research would Advance Research Planning not be appropriate for? Tick all that apply | 1) Observational studies that do not involve invasive procedures (e.g studies collecting clinical/routine data or involving interviews/ethnography) 2) Observational studies that involve procedures (e.g blood tests, scans, wearable technology) 3) Interventional studies not involving medicinal products (non-CTIMPs) 4) Interventional studies involving medicinal products (e.g CTIMPs, ATIMPs) 5) Other (*please specify*) |

| **Q3. In your view, which groups of people might Advance Research Planning be most appropriate for?** | 1) People living with conditions such as dementia, Parkinson’s, or Huntington’s disease 2) People at high risk of developing conditions such as dementia, Parkinson’s, or Huntington’s disease 3) Older people 4) People at risk of acute medical events such as stroke 5) Other (*please specify*) |
| --- | --- |
| Which groups of people would Advance Research Planning not be appropriate for? Tick all that apply | 1) People living with conditions such as dementia, Parkinson’s, or Huntington’s disease 2) People at high risk of developing conditions such as dementia, Parkinson’s, or Huntington’s disease 3) Older people 4) People at risk of acute medical events such as stroke 5) Not appropriate for any group 6) Other (*please specify*) |

| **Q4. Advance Research Planning can be undertaken at different times. Please indicate the extent to which you agree with the following statements:** | Strongly  agree | Agree | Unsure | Disagree | Strongly  disagree |
| --- | --- | --- | --- | --- | --- |
| Advance Research Planning should be undertaken at the same time as **other planning processes** (e.g when having advance care planning discussions or making Power of Attorney arrangements) | 1 | 2 | 3 | 4 | 5 |
| Advance Research Planning should be undertaken after someone is **diagnosed** as having (or being at risk of) a condition that might affect their capacity in the future (e.g dementia, stroke, approaching the end of life) | 1 | 2 | 3 | 4 | 5 |
| Advance Research Planning should be undertaken when someone is being approached about a **specific study** they may wish to participate in (e.g when joining a research registry, entering an initial observational stage of an interventional study) | 1 | 2 | 3 | 4 | 5 |
| Advance Research Planning should be undertaken at **opportunistic or ad hoc** times (e.g at any point when motivated or interested in doing so, as with decisions about organ donation) | 1 | 2 | 3 | 4 | 5 |
| Advance Research Planning should be **reviewed** at different timepoints (e.g at regular timepoints or when there are changes in their personal circumstances or clinical condition) | 1 | 2 | 3 | 4 | 5 |

| **Q5. Advance Research Planning can cover different content. Please indicate the extent to which you agree with the following statements:** | Strongly  agree | Agree | Unsure | Disagree | Strongly  disagree |
| --- | --- | --- | --- | --- | --- |
| Advance Research Planning should include nominating **who** makes decisions on their behalf (e.g naming a particular person such as their spouse/partner) | 1 | 2 | 3 | 4 | 5 |
| Advance Research Planning should include their **general wishes** about research they would or would not want to participate in (e.g studies related to their condition only or any study they are eligible for) | 1 | 2 | 3 | 4 | 5 |
| Advance Research Planning should include their wishes about what **specific types of research** they would or would not want to participate in (e.g observational or interventional, anticipated benefits and risks involved, involving specific procedures such as blood tests or scans) | 1 | 2 | 3 | 4 | 5 |

| **Q6. Advance Research Planning can be used be used to inform a decision about whether someone who lacks capacity should take part in a research study in different ways. Please indicate the extent to which you agree with the following statements:** | Strongly  agree | Agree | Unsure | Disagree | Strongly  disagree |
| --- | --- | --- | --- | --- | --- |
| Wishes expressed through Advance Research Planning should be considered **legally binding** (they should be followed regardless of what others involved in the process think) | 1 | 2 | 3 | 4 | 5 |
| Wishes expressed through Advance Research Planning should be considered **directive** (they should directly inform the decision, although they do not have to be followed if there are reasonable views otherwise) | 1 | 2 | 3 | 4 | 5 |
| Wishes expressed through Advance Research Planning should be considered **advisory** (they can help or contribute to the decision) | 1 | 2 | 3 | 4 | 5 |

| **Q7. Advance Research Planning can take a number of** **different forms. Please indicate the extent to which you agree with the following statements:** | Strongly  agree | Agree | Unsure | Disagree | Strongly  disagree |
| --- | --- | --- | --- | --- | --- |
| Advance Research Planning should be an **informal discussion** which is then written down by the individual themselves and/or the professional involved (e.g the discussion is summarised by a member of their clinical team or care provider in their notes) | 1 | 2 | 3 | 4 | 5 |
| Advance Research Planning should be a **formal process** which is documented by the individual and a formal copy shared with others (e.g similar to an advance statement about wishes and care preferences) | 1 | 2 | 3 | 4 | 5 |
| Advance Research Planning should be a **legal process** in which wishes about research are documented in a legal document which is then registered (e.g similar to the process for Power of Attorney) | 1 | 2 | 3 | 4 | 5 |

| **Q8. Different people could be involved in the process of Advance Research Planning Please indicate the extent to which you agree with the following statements:** | Strongly  agree | Agree | Unsure | Disagree | Strongly  disagree |
| --- | --- | --- | --- | --- | --- |
| The person who is involved in making **decisions** with or for the person should be involved (example: spouse or adult child, someone with Power of Attorney) | 1 | 2 | 3 | 4 | 5 |
| A doctor or other health professional who is **part** **of their healthcare team** should be involved | 1 | 2 | 3 | 4 | 5 |
| A doctor or other research professional who is **part of a research team** should be involved | 1 | 2 | 3 | 4 | 5 |

**Q9. What might the barriers to undertaking Advance Research Planning be, and what could help support it?**

**Q10. Do you have any other comments about Advance Research Planning?**

### **Part C. Contact about next stage of the project**

**If you are willing to be contacted about taking part in an optional interview to talk about your views about advance research planning, please provide your contact details below.**

Name:

Email address:

Phone no:

Preferred form of contact: phone/email

Preferred time to be contacted: morning/afternoon/evening/anytime

**Thank you**

Thank you for taking part in our survey.

If you want to get in touch with us further, please contact: Dr Victoria Shepherd at Cardiff University 02920687641
