## Appendix 2. CONSULT-ADVANCE study questionnaire for members of the public for "Planning ahead for research participation: survey of public and professional stakeholders’ views about the acceptability and feasibility of advance research planning"

### **Part A. About you**

**Which group best describes your main interest in this area?**

- Personal experience of living with a condition that may affect my memory and understanding
- Family member/friend of someone living with a condition that may affect their memory and understanding
- Member of the public interested in this topic
- Other (*please specify*)

**Is your main interest in this area in relation to?**

- Dementia
- Stroke
- Parkinsons disease
- Huntington’s disease
- Palliative care or end of life care
- Other (*please specify*)

**Where do you usually live?**

- England
- Northern Ireland
- Scotland
- Wales
- Other (*please specify*)

**What is your current gender identity?**

- Female
- Male
- Other (*please specify*)

**What is your age range?**

- 18-24
- 25-34
- 35-49
- 50-64
- 65+

**What is your ethnic group?**

(Choose one option that best describes your ethnic group or background)

- White
- Mixed / Multiple ethnic groups
- Asian / Asian British
- Black / African / Caribbean / Black British
- Other ethnic group

**Have you previously taken part in research, for example into a condition that affects memory and understanding?**

- Yes I have previously taken part in research
- No I have not previously taken part in research
- Other (*please specifiy*)

**Have you previously taken part in any advance planning activities for either you or someone else (****e.g setting up Power of Attorney, creating an advance directive or statement about future care)?**

- Yes I have previously taken part in advance planning activities
- No I have not previously taken part in advance planning activities
- Other (*please specifiy*)

### **Part B. Your views about research**

**Imagine the following situation:**

You have dementia or another condition and you have quite a few troubles with memory, thinking and making decisions.

You could take part in a research study. This study is testing ways to improve care or treatments for people with your condition.

An independent ethics committee has reviewed the study to make sure it is safe and ethical and there are arrangements in place to ensure your privacy will be protected.

At this point in time, you are not able to make your own decision about taking part in the study.

**How much do you agree or disagree with the following statements (remember the questions are hypothetical):**

| **Q1. I would be willing to be included in a research study that involves:** | **Strongly agree** | **Agree** | **Neither disagree nor agree** | **Disagree** | | **Strongly disagree** |
| --- | --- | --- | --- | --- | --- | --- |
| a) Asking me questions in a survey or interview *(e.g asking about my experiences or opinions)* | 1 | 2 | 3 | 4 | 5 | |
| b) Observing my behaviour *(e.g watching how I act if I listen to music)* | 1 | 2 | 3 | 4 | 5 | |
| c) Testing my memory or thinking *(e.g asking me to draw a picture or remember specific words)* | 1 | 2 | 3 | 4 | 5 | |
| d) Giving me psychological therapy *(e.g counselling for anxiety or depression)* | 1 | 2 | 3 | 4 | 5 | |
| e) Giving me physiotherapy *(e.g moving my arms or legs, massaging my muscles)* | 1 | 2 | 3 | 4 | 5 | |
| f) Giving me experimental medicine *(e.g a drug that might help my condition)* | 1 | 2 | 3 | 4 | 5 | |
| g) Taking x-rays or scans of my body *(e.g to help researchers see how my condition is affecting my brain)* | 1 | 2 | 3 | 4 | 5 | |
| h) Taking measurements about my body *(e.g my weight, blood pressure)* | 1 | 2 | 3 | 4 | 5 | |
| i) Putting something on my body, like a bracelet, that keeps track of information *(e.g how much time I spend in bed)* | 1 | 2 | 3 | 4 | 5 | |
| j) Taking a sample of my blood or other body fluid for **genetic research** *(e.g to find out if I and my relatives have a gene that increases the risk of getting dementia)* | 1 | 2 | 3 | 4 | 5 | |
| k) Taking a sample of my blood or other body fluid for **non-genetic research** *(e.g to find out if my blood shows I had an infection in the past that increases my risk of a condition*) | 1 | 2 | 3 | 4 | 5 | |
| l) Looking at my personal records, such as medical records or test results held at my GP practice or hospital *(e.g to study how a past illnesses might be related to my condition)* | 1 | 2 | 3 | 4 | 5 | |
| m) Accessing stored samples of my blood, body fluids or other tissues *(e.g If I had blood taken in the past for another reason, researchers might ask the hospital for access to that blood for study)* | 1 | 2 | 3 | 4 | 5 | |

| **Q2. I would be willing to be included in a research study that:** | Strongly agree | Agree | Neither agree nor disagree | Disagree | Strongly disagree |
| --- | --- | --- | --- | --- | --- |
| Benefits me directly (e.g taking part in research could improve my quality of life) | 1 | 2 | 3 | 4 | 5 |
| May not benefit me directly but could help other people with my condition | 1 | 2 | 3 | 4 | 5 |
| May not have benefits for me or other people with my condition, but could help researchers understand other diseases or health problems | 1 | 2 | 3 | 4 | 5 |

### **Part C. Your views about advance research planning**

**Advance Research Planning** is a process where you can express your wishes about being involved in research studies in the future.

You take part in the process of Advance Research Planning at a time when you are able to think through your options and make choices.

It might include writing down **what your wishes are** about being involved in different types of research, and **who you would like to be involved** in making a decision on your behalf.

If you later lose the ability to make decisions due to a medical condition, Advance Research Planning will tell people what your wishes are, such as your doctor, a family member or friend, or a researcher.

| **Q3. If it were possible for you to take part in Advance Research Planning, how interested would you be in doing this?** | 1) Very interested *(skip next question)* 2) Somewhat interested *(skip next question)* 3) Unsure *(skip next question)* 4) Not very interested 5) Not at all interested |
| --- | --- |
| Why are you not interested in taking part in Advance Research Planning? | 6) I am not interested in taking part in research in the future 7) I do not think it is important to express my wishes for taking part in future research 8) I would prefer for someone else to make decisions about my participation in research if I am no longer able to make my own decisions 9) I do not think it matters what happens after I lose the ability to make decisions 10) I am not sure 11) Other (please specify) |

**If you are willing to be contacted about taking part in an optional interview to talk about your views about advance research planning, please provide your contact details below.**

**I am willing to be contacted by researchers from Cardiff University about taking part in an optional interview:**

Name:

Email address:

Phone no:

Preferred form of contact: phone/email

Preferred time to be contacted: moring/afternoon/eveneing/anytime

**Thank you**

Thank you for taking part in our survey.

If you want to get in touch with us further, please contact: Dr Victoria Shepherd at Cardiff University
