## Supplementary File 1 CONSULT-ADVANCE Study Protocol v1.0 03.11.2022 for "Planning ahead for research participation: survey of public and professional stakeholders’ views about the acceptability and feasibility of advance research planning"

PARTICIPATION: STAKEHOLDERS' VIEWS

ABOUT ADVANCE RESEARCH PLANNING

(CONSULT-ADVANCE STUDY)

PROTOCOL V1.0 03.11.2022

| | |
|---|---|
| **Funder:** | **CONSULT project funded by Health and Care Research Wales** |
| **REC ref:** | **TBC** |
| **Q-Pulse Document Template Number :** | **TPL/003/2** |

**SIGNATURE PAGE**

The undersigned confirm that the following protocol has been agreed and accepted and that the Chief Investigator agrees to conduct the study in compliance with the approved protocol and will adhere to the principles outlined in the relevant regulations, GCP guidelines, and CTR's SOPs.

I agree to ensure that the confidential information contained in this document will not be used for any other purpose other than the evaluation or conduct of the study.

I also confirm that I will make the findings of the study publicly available through publication or other dissemination tools without any unnecessary delay and that an honest accurate and transparent account of the study will be given; and that any discrepancies from the study as planned in this protocol will be explained.

| | | |
|---|---|---|
| **Director:** | | |
| **Name** | **Signature** | **Date** |
| **Chief Investigator:** | | |
| **Name** | **Signature** | **Date** |

**General Information** This protocol describes the CONSULT-ADVANCE Study and provides information about the procedures for entering participants into the study. The protocol should not be used as a guide, or as an aide-memoire for the treatment of other participants. Every care has been taken in drafting this protocol; however, corrections or amendments may be necessary. These will be circulated to the known Investigators in the study. Problems relating to the study should be referred, in the first instance, to CTR.

**Contact details – Chief Investigator/s & Co-Investigator/s**

CHIEF INVESTIGATOR

Dr Victoria Shepherd

Research Fellow

Centre for Trials Research, Cardiff University

4th floor Neuadd Meirionnydd, Heath Park, Cardiff

CF14 4YS

CO-INVESTIGATOR(S)

Prof Kerry Hood

Director of Centre for Trials Research

Dr Fiona Wood

Professor of Medical Sociology, Division of Population Medicine

**Glossary of abbreviations**

| | |
|---|---|
| **ACP** | Advance care planning |
| **ARP** | Advance research planning |
| **CF** | Consent Form |
| **CI** | Chief Investigator |
| **CTR** | Centre for Trials Research |
| **CTU** | Clinical Trials Unit |
| **CU** | Cardiff University |
| **GCP** | Good Clinical Practice |
| **HRA** | Health Research Authority |
| **IC** | Informed consent |
| **ICH** | International Conference on Harmonization |
| **NIHR** | National Institute for Health and Care Research |
| **REC** | Research Ethics Committee |
| **SOP** | Standard Operating Procedure |

### 1 Amendment History

The following amendments and/or administrative changes have been made to this protocol since the implementation of the first approved version.

| Amendment No. | Protocol version no. | Date issued | Summary of changes made since previous version |
|---|---|---|---|

### 2 Synopsis

| | |
|---|---|
| **Short title** | Planning ahead for research participation: stakeholders' views about advance research planning |
| **Acronym** | CONSULT-ADVANCE Study |
| **Internal ref. no.** | Sub study of UID 1010 |
| **Funder and ref.** | NIHR Advanced Fellowship (CONSULT) funded by Health and Care Research Wales |
| **Study design** | Survey |
| **Study participants** | Public stakeholders including people living with dementia or another condition that may affect their memory and understanding or a family member/friend; professional stakeholders including researchers interested in dementia or other capacity-affecting conditions or those interested in the ethical and practical aspects of advance planning |
| **Planned sample size** | Approx. 150 |
| **Inclusion criteria** | • People with personal experience of either living with dementia or another condition which may affect their ability to consent to research, a family member or friend of someone with such a condition, or a member of the public interested in research or advance planning<br>OR<br>• Researcher or other professional with an interest in dementia or another capacity-affecting condition, or an interest in advance planning |
| **Exclusion criteria** | • Is unable to understand English sufficiently to comprehend the study information and complete the survey in English |
| **Planned study period** | 6 months |
| **Objectives** | 1) Explore stakeholders' views about the acceptability of planning for research participation during future periods of impaired capacity<br>2) Explore stakeholders' views about the feasibility of advance research planning<br>3) Identify barriers and facilitators to the implementation of advance research planning |

### 3 Study lay summary

Around 2 million people in the UK have significantly impaired decision-making. Over the coming decades, this number is likely to grow considerably due to an ageing population and a rise in conditions such as dementia. Research is essential in order to improve evidence -based care for these populations who often have the most complex care needs. However, research involving people whose capacity to consent is impaired raises a number of ethical and practical issues.

Involving people with conditions such as dementia in research may require asking a family member to decide on their behalf whether they should take part or not, but some family members do not know what the person's wishes and preferences about research would be and find it difficult to make a decision.

There has been a growing focus on advance care planning that provides people with the opportunity to discuss their future preferences about treatment and care should they be unable to decide for themselves. This includes arrangements to appoint a Power of Attorney and aims to ensure that decisions are made in line with those preferences and by the person who knows them best. However, in the UK these legal arrangements do not include decisions about research, despite 'Advance Research Directives' being introduced in other countries such as US and Canada. Extending these arrangements to include research preferences may support peoples' autonomy through providing an opportunity to express their wishes about future research participation and who makes a decision about research on their behalf.

Further work is needed to explore the views of people living with conditions such as dementia, their families, and researchers and other professional groups about the acceptability of planning for future research participation in the UK. We also need to explore whether advance research planning is feasible, and what factors might affect the use of advance research planning in practice.

We will conduct an online survey with people living with dementia or other conditions which can affect memory and understanding, family members and other members of the public, and with researchers and other professionals to ask for their views about advance research planning. We will use the findings to plan future research and to develop an advance research planning intervention.

### 4 Background

More than 920,000 people in the UK are living with dementia, and this is expected to rise to over a million by 2024 [1]. Alongside a range of other conditions and disabilities, dementia contributes to the approximately 2 million people in the UK who have significantly impaired decision-making [2]. Conducting research with people who have significantly impaired decision-making relies on proxy decision-makers to make decisions about participation on their behalf. In the UK, proxy decisions made on behalf of people who lack capacity to consent should be based on what the person's wishes and feelings about taking part in the study would be [3, 4]. However, people rarely discuss their preferences about research and so it can be difficult for families to make a decision about whether their relative should participate in a research study, and many experience an emotional and decisional burden as a result [5]. Previous studies suggest that proxies' views often differ from what the potential participants would want [6], including a tendency to underestimate the willingness of older adults to participate in research [7]. Knowing the person's wishes may help families or, in the event there is no-one available to act in a personal capacity, a member of their care team, acting as consultees or legal representatives in the event of a loss of capacity [5]. It may also contribute to better inclusion in trials of people with impaired capacity to consent as this group is frequently excluded from trials [8] – primarily due to the ethical and legal complexities around consent [9]. People with impaired capacity to consent have been recognised by the NIHR INCLUDE initiative as being as an under-served group in research [10].

##### *Use of advance care planning for decisions about care*

There is an increasing focus on supporting people living with conditions such as dementia and cancer to discuss their preferences about future care and treatment options prior to any loss of capacity. Advance care planning (ACP) is viewed as a way of as enabling people with capacity to think about the meanings and consequences of different future scenarios and discuss their goals and preferences with family members and their healthcare providers [11]. It encourages them to identify a proxy decision-maker and to record and regularly review any preferences, so that their preferences can be taken into account should they, at some point, be unable to make their own decisions [11]. ACP forms part of NICE guidelines for end-of-life care for adults [12], dementia [13], and decision-making and mental capacity [14] and they have produced a website of resources on advance care planning for social care [15]. In England and Wales, under the Mental Capacity Act 2005 here are legal provisions for making advance decisions to refuse treatment for (physical health) conditions and for appointing someone to make decisions about health and care through Lasting Power of Attorney arrangements [4]. Reforming the Mental Health Act 1983 to include equitable provision for advance decision-making (ADM) in mental health has been the subject of recent consultation [16]. There are also prior arrangements for donating brain and other tissue for research after death [17]. However, despite the need to embed research into care pathways being a key strategic aim in the UK [18], advance care planning discussions and formal processes such as Power of Attorney arrangements do not currently extend to preferences and proxy decisions about research participation in the UK.

##### *Advance research planning in other countries*

Other jurisdictions have established processes for planning ahead for research participation in the event of a loss of capacity. This includes the US and in Canada, where more advance research planning, including the ability to make advance research directives (ARDs) which direct proxies' decision-making rather than advise, was introduced several decades ago [19]. Since 2007, Australia's National Statement on Ethical Conduct in Human Research has enabled a formal process for researchers to discuss and document views on future research participation with participants who anticipate periods of cognitive impairment [20]. More recently, there have been moves to integrate ARDs into the European legal framework [21], with legislation awaiting implementation in Switzerland [19]. Germany has gone one stage further with recent changes in legislation to require an ARD to be in place in order for people lacking capacity to consent to be included in research with group benefit [22]. Advance research planning (ARP) has been proposed as a means to overcome the challenges associated with proxy decision-making for research [23], and has been explored in populations including older people [6, 24], people living with dementia [22, 25], care home residents [26], palliative care [27], as well as acute conditions such as stroke [28].

It is thought that ARP can promote respect for the autonomy and self-determination of people who wish to plan for future incapacity, help ensure that proxy decision-making reflects the values and wishes of those lacking capacity to consent, and support the inclusion in research of those who have expressed their prior interest in participation [29]. It may also reduce the burden experienced by proxies when making a decision on their behalf [30]. Studies in other jurisdictions show widespread support for various forms of ARP, including Australian studies involving older people [23] and dementia researchers [25], and . ARP has support from organisations such as Alzheimer Europe who encourage the use of advance directives to record peoples' wishes to participate (or not participate) in research as it respects their right to self-determination and their desire to do something constructive which may eventually benefit others with a similar medical condition [31].

##### *Uncertainties about advance research planning in the UK*

In the UK, family members with experience of acting as research proxy are supportive of ARP, including the introduction of a process to nominate someone to make future decisions about research, with some proxies suggesting that there were benefits to extending the existing arrangements for LPA for health and welfare [32]. The Nuffield Council on Bioethics has long recommended the commissioning of research on the feasibility of developing a (non-binding) advance statement on research participation which could influence decisions on research participation after loss of capacity (Recommendation 18) [33].

However, ARP raises a number of ethical and practical concerns that can be summarised as 'what' (what types of research should ARP be considered for, what information should ARP include and in what format), 'when' (when is the best time for public or patients to undertake ARP and when is the best time to convey that information to others), 'who' (which groups of public/patients should be approached to undertake ARP and which groups of professionals should be involved), and 'how' (how should ARP be implemented in practice and which kind of safeguards might need to be in place) [24]. There are legal uncertainties about how (or if) it would align with mental capacity legislation across the UK (e.g Mental Capacity Act 2005 in England and Wales [4], Adults with Incapacity (Scotland) Act 2000 [34]) and clinical trials regulation (e.g Medicines for Human Use (Clinical Trials) Regulation [3]). There is also the risk that introducing the opportunity to express future wishes about research participation via ARP may also lead to this becoming a legal requirement in order to involve people who lack capacity to consent (as is the case for some types of research in Germany [22]) which would disadvantage people who are unable to or do not wish to take part in ARP, which would particularly exclude groups such as people with a learning disability and those least likely to have family members to involve in the process.

There is also wide variation in the terminology and definitions used in the literature and ethical and legal frameworks to describe the process of advance planning for research participation, and the degree to which it can be considered legally binding. For the purposes of this study, ARP is viewed as conceptually different to advance consent (or 'advanced consent') where ARP is considered a mechanism for people to express their preferences about the *general* aims, methods, risks and burdens, and types of research studies they would wish to participate in, rather than *specific* studies [24]. However, views about the acceptability and feasibility of ARP will include views about the type/level of information needed in order to express a preference about participation, previously described as either 'type disclosure' which requires general information about the type of research or 'token disclosure' which requires details of a specific trial that may not be available in advance [19]).

Despite introduction of ARP in other jurisdictions, no studies have explored stakeholders' views about ARP in the UK. This survey will explore a range of public and professional stakeholders' views about the acceptability and feasibility of ARP in the UK (including the degree to which preferences and wishes should be considered binding) and identify the barriers and facilitators to implementing ARP. The findings will be used to support the development of interventions to support people to express their future wishes about research through processes such as ARP.

### 5 Study objectives

The objectives of this study are to:

1) Explore stakeholders' views about the acceptability of planning for research participation during future periods of impaired capacity
2) Explore stakeholders' views about the feasibility of advance research planning
3) Identify barriers and facilitators to the implementation of advance research planning

### 6 Study design and setting

This exploratory study will use an online survey to explore attitudes towards advance research planning and its acceptability to members of the public and professionals involved in research. Members of the public with personal experience of conditions that may affect capacity to consent will be recruited, alongside researchers and other professionals with an interest in research into capacity-affecting conditions.

Participants will be invited to participate in an online survey, with two versions of the survey (one for public stakeholders and one for professionals) which have been designed to capture the different, although corresponding, perspectives of the two groups. The study will commence in November 2022 with a duration of 6 months.

#### 6.1 Risk assessment

A comprehensive Risk Assessment (RA) for the study is in place, following Centre for Trials Research Standard Operating Procedures for RA. This details any risks/potential risks of the study, including any potential risk/harm to participants and/or researchers. The RA will contain a plan for minimising potential risks/harms and managing risks/harms should they occur.

This study has been categorised as a low risk, where the level of risk is 'comparable to the risk of standard care'. A copy of the risk assessments may be requested from the CI.

### 7 Participant selection

The aim of the survey is to obtain the views and perspectives of a wide range of public and professional stakeholders. This includes people living with a variety of conditions that may affect their ability to provide consent to participate in research, family members of someone living with a potentially capacity-affecting condition, and researchers and other professionals with an interest in research into capacity affecting conditions.

A potential participant will be eligible if they meet all the inclusion criteria. A potential participant will not be eligible if any of the exclusion criteria apply.

##### 7.1 Inclusion criteria

- People with personal experience of either living with dementia or another condition which may affect their ability to consent to research, a family member or friend of someone with such a condition, or a member of the public interested in research or advance planning
  OR
- Researcher or other professional with an interest in dementia or another capacity-affecting condition, or an interest in advance planning

##### 7.2 Exclusion criteria

- Is unable to understand English sufficiently to comprehend the study information and complete the survey in English

#### 8 Recruitment, Screening and registration

##### 8.1 Participant identification

There are two types of stakeholder groups who will be participants in this study.

We will be recruiting public stakeholders (family members of someone with dementia or another impairing condition, people living with condition such as dementia, or people interested in research into conditions such as dementia) through a number of routes. Potential participants will be identified through social media platforms (e.g Twitter), research networks (e.g Alzheimer's Society Volunteer Research Network), and community or charity groups (e.g dementia support groups). We will also be recruiting through Join Dementia Research (JDR). This is an online registry that enables volunteers with memory problems or dementia, carers of those with memory problems or dementia and healthy volunteers to sign up and register their interest in taking part in research. The purpose of JDR is to allow such volunteers to be identified by researchers as potentially eligible for their studies using a set of 'matching criteria'. Researchers who register their study with JDR will see a list of matched potential volunteers and can then contact volunteers, in line with the volunteers' preferred method of contact, to further discuss potential inclusion. JDR is funded by Department of Health working in partnership with the charities Alzheimer Scotland, Alzheimer's Research UK and Alzheimer's Society and is Health Research Authority (HRA) endorsed. The online service and all associated documentation, methods of contacting volunteers and handling of data, were reviewed by a specially convened HRA committee which included experts in research ethics, data protection and information governance. Formal endorsement was issued by the HRA in a letter dated 5 November 2019. There is an existing Data Processing Agreement in place between Cardiff University and JDR covering data protection and confidentiality.

We will be recruiting researchers and other professional stakeholders with an interest in dementia or another condition which may affect their capacity to consent to research in a number of ways. Potential participants will be identified through social media platforms (e.g Twitter) and research networks (e.g MRC-NIHR Trials Methodology Research Partnership, British Society for Gerontology). We will also search databases of current and previously funded studies (e.g NIHR portfolio) to identify researchers who are working in relevant areas and recruit other professional stakeholders such as research ethics committee members through organisations such as the HRA.

Potential participants will be provided with a link to access the online survey that is relevant to the stakeholder group they identify with (public or professional). Each survey will contain a home page with participant information about the study, followed by the questionnaire pages.

##### 8.2 Informed consent

The study will be conducted using an online survey tool (Qualtrics). Following the guidance from the Health Research Authority on conducting online surveys, no separate Participant Information Sheet or consent form will be provided [35]. The opening page for the survey will provide a brief introduction of the aim of the study, how the information collected will be used and stored, and how the findings will be used. Participants will be asked to tick a box at the start of the survey to confirm that they agree to participate. Participants who do not confirm will be considered as having withdrawn from further participation.

This project will comply with GCP and Centre for Trials Research SOPs and participants' confidentiality will be maintained. The online survey tool is GDPR compliant. All data will be kept for for no less than end of project + 5 years or at least 2 years post publication in line with Cardiff University's Research Governance Framework Regulations for clinical research. The data will be stored confidentially on password protected servers maintained on the Cardiff University Network and in accordance with the General Data Protection Regulations (2016)(GDPR).

#### 9 Withdrawal

Participants will be informed that they are able to withdraw from the study, should they wish to do so, by contacting the CI. Participants will be completing a questionnaire at a single timepoint and will be providing anonymous responses therefore data already provided cannot be withdrawn from the analysis.

### 10 Data collection

Two surveys have been developed based on previous surveys conducted with public and professional stakeholders in other countries (e.g [23, 25]) and adapted for the UK context and the aims of this study. The survey questions and information pages have been reviewed by a lay advisory group and piloted prior to use.

Data collection will be via an online survey tool (Qualtrics) which is widely used for similar surveys and for which CTR has a licence. Participants will be provided with a survey link URL. The opening page for the survey will provide a brief introduction of the aim of the study. Participants who agree to participate will be asked to register their details online, including specifying which stakeholder group they consider themselves to be associated with. Participants will be asked to tick a box at the start of the survey to confirm that they consent to participate. Participants who do not confirm will be considered as having withdrawn from further participation.

### 11 Statistical considerations

As an exploratory survey, there is no sample size calculation. However, an estimated sample size of approx. 150 participants is informed by similar surveys [36].

### 12 Analysis

The data will be analysed using Qualtrics online survey software. Participant characteristics will be presented descriptively. Descriptive statistics will be used to summarise the survey responses.

### 13 Data Management

This project will comply with GCP and Centre for Trials Research SOPs and participants' confidentiality will be maintained. The online survey tool is GDPR compliant. All data will be kept in line with Cardiff University's Research Governance Framework Regulations for non-clinical research for no less than end of project + 5 years or at least 2 years post publication. The data will be stored confidentially on password protected servers maintained on the Cardiff University Network server in accordance with the GDPR regulations and only accessed by named individuals within the research team. The data controller is Cardiff University. The legal basis for processing participant data is as a public task and on the basis of public interest.

### 14 Protocol/GCP non-compliance

The Chief Investigator will report any non-compliance to the trial protocol or the conditions and principles of Good Clinical Practice to the CTR in writing as soon as they become aware of it.

### 15 Regulatory Considerations

#### 15.1 Ethical and governance approval

The protocol, participant information and questionnaire, and any proposed advertising material will be submitted for written approval to an appropriate Research Ethics Committee (REC) prior to the start of the study taking place. As NHS patients or users of the NHS, or their relatives, will not be involved in the study, a favourable ethical opinion will be sought from the School of Medicine Research Ethics Committee, Cardiff University. Approval will also be sought regarding any amendments to the study.

The study will be conducted in accordance with ICH GCP requirements, and in accordance with the recommendations for research on human participants adopted by the 18th World Medical Assembly, Helsinki 1964 as amended. The Health Research Authority guidance on consent and participant information sheets and online methods of 'eConsent' will be followed as appropriate. Participants will be provided with sufficient information prior to any decision whether to participate and informed that it is voluntary.

#### 15.2 Indemnity

Cardiff University will provide indemnity and compensation in the event of a claim by, or on behalf of participants, for negligent harm as a result of the study design and/or in respect of the protocol authors/research team. Cardiff University does not provide compensation for non-negligent harm.

#### 15.3 Funding

This study is funded through an NIHR Advanced Fellowship funded by Health and Care Research Wales.

#### 16 Publication policy

The study results will be published in peer-reviewed academic journals, and the results presented at conferences, and through other dissemination events or outputs. Participants will not be providing contact details therefore we will not be able to share details of the findings directly, but summaries of the study findings and information about how to access a copy of the full results (e.g. online journal article) will be shared via the project website. The Lay Advisory Group who support the project will be asked for their assistance in ensuring the material prepared for the participants and general public is accessible and appropriate.

The funders will have no role in decisions on publication. The funding source and other support will be acknowledged.

2. Care Quality Commission. Monitoring the use of the mental Capacity Act Deprivation of Liberty Safeguards in 2013/14. 2015.

3. The Medicines for Human Use (Clinical Trials) Regulations 2004 SI No.1031. 2004.

4. HMSO, London. Mental Capacity Act 2005. 2005.

12. NICE Guideline [NG142] End of life care for adults: service delivery. 2019.

13. National Institute for Health and Care Excellence. NICE guideline NG97 Dementia: assessment, management and support for people living with dementia and their carers. 2018.

14. National Institute for Health and Care Excellence. NICE guideline NG108 Decision-making and mental capacity. NICE; 2018.

15. Advance care planning for NICE Communities, social care. NICE. https://www.nice.org.uk/about/nice-communities/social-care/quick-guides/advance-care-planning. Accessed 17 Oct 2022.

16. Owen GS, Gergel T, Stephenson LA, Hussain O, Rifkin L, Keene AR. Advance decision-making in mental health – Suggestions for legal reform in England and Wales. International Journal of Law and Psychiatry. 2019;64:162–77.

17. Body, organ and tissue donation. HTA. https://www.hta.gov.uk/guidance-public/body-organ-and-tissue-donation. Accessed 20 Oct 2022.

18. National Institute for Health Research,. Best Research for Best Health: The Next Chapter. 2021.

19. Heinrichs B. Advance research directives: avoiding double standards. BMC Medical Ethics. 2021;22:137.

20. National Statement on Ethical Conduct in Human Research (2007) - Updated 2018 | NHMRC. 2018.

21. Andorno R, Gennet E, Elger B, Jongsma K. Integrating Advance Research Directives into the European Legal Framework. European Journal of Health Law. 2016;23:158–73.

22. Jongsma K, Perry J, Schicktanz S, Radenbach K. Motivations for people with cognitive impairment to complete an advance research directive – a qualitative interview study. BMC Psychiatry. 2020;20:360.

23. Ries NM, Mansfield E, Sanson-Fisher R. Advance Research Directives: Legal and Ethical Issues and Insights from a National Survey of Dementia Researchers in Australia. Medical Law Review. 2020;28:375–400.

24. Werner P, Schicktanz S. Practical and Ethical Aspects of Advance Research Directives for Research on Healthy Aging: German and Israeli Professionals' Perspectives. Front Med (Lausanne). 2018;5.

25. Ries N, Mansfield E. Advance Research Directives: Dementia Researchers' Views on a Prototype Directive and Implementation Strategies. Ethics Hum Res. 2021;43:10–25.

26. Wood F, Prout H, Bayer A, Duncan D, Nuttall J, Hood K, et al. Consent, including advanced consent, of older adults to research in care homes: a qualitative study of stakeholders' views in South Wales. Trials. 2013. https://doi.org/10.1186/1745-6215-14-247.

27. Stanley S, Nwosu AC. Case Report: The use of advanced consent methodology and healthcare professional consultee to facilitate research participation in dying patients. 2021.

28. Shamy M, Dewar B, Niznick N, Nicholls S, Dowlatshahi D. Advanced consent for acute stroke trials. The Lancet Neurology. 2021;20:170.

29. Pierce R. A changing landscape for advance directives in dementia research. Social Science & Medicine. 2010;70:623–30.

30. Burns KEA, Prats CJ, Maione M, Lanceta M, Zubrinich C, Jeffs L, et al. The experience of surrogate decision makers on being approached for consent for patient participation in research: A multicenter study. Annals of the American Thoracic Society. 2017;14:238–45.

31. Advance Directives - Alzheimer Europe Position Paper 2005. Alzheimer Europe; 2005.

32. Shepherd V, Griffith R, Hood K, Sheehan M, Wood F. "There's more to life than money and health": Family caregivers' views on the role of Power of Attorney in proxy decisions about research participation for people living with dementia. Dementia (London). 2019;:1471301219884426.

33. Nuffield Council on Bioethics. Dementia: ethical issues. 2009.

34. Adults with Incapacity (Scotland) Act 2000. Statute Law Database; 2000.

35. Health Research Authority,, Medicines and Healthcare products Regulatory Agency,. HRA and MHRA Joint statement on seeking consent by electronic methods (eConsent). 2018.

36. Ries N, Mansfield E, Sanson-Fisher R. Planning Ahead for Dementia Research Participation: Insights from a Survey of Older Australians and Implications for Ethics, Law and Practice. J Bioeth Inq. 2019;16:415–29.
