## Supplementary File 2 Additional survey data for "Planning ahead for research participation: survey of public and professional stakeholders’ views about the acceptability and feasibility of advance research planning"

*Figure 1. Public stakeholders’ views about timing of advance research planning*

|  | Strongly agree | Agree | Neither agree nor disagree | Disagree | Strongly disagree |
| --- | --- | --- | --- | --- | --- |
| Same time as ACP | 117 | 119 | 30 | 4 | 1 |
| After diagnosis | 105 | 73 | 49 | 41 | 3 |
| When approached for a study | 83 | 110 | 59 | 15 | 3 |
| Opportunistic or ad hoc | 85 | 95 | 67 | 18 | 6 |
| Reviewed at different timepoints | 123 | 123 | 20 | 2 | 3 |

*Figure 2. Professional stakeholders’ views about timing of advance research planning*

|  | Strongly agree | Agree | Neither agree nor disagree | Disagree | Strongly disagree |
| --- | --- | --- | --- | --- | --- |
| Same time as ACP | 16 | 23 | 6 | 1 | 0 |
| After diagnosis | 12 | 19 | 12 | 2 | 1 |
| When approached for a study | 11 | 22 | 7 | 6 | 0 |
| Opportunistic or ad hoc | 7 | 22 | 13 | 3 | 1 |
| Reviewed at different timepoints | 21 | 19 | 4 | 2 | 0 |

*Figure 3. Public stakeholders’ views about the content of advance research planning*

|  | Nominating who | General wishes | specific types |
| --- | --- | --- | --- |
| Strongly agree | 163 | 159 | 164 |
| Agree | 92 | 95 | 93 |
| Neither agree nor disagree | 12 | 11 | 10 |
| Disagree | 1 | 2 | 0 |
| Strongly disagree | 1 | 2 | 2 |

*Figure 4. Professional stakeholders’ views about the content of advance research planning*

|  | Nominating who | General wishes | Specific studies |
| --- | --- | --- | --- |
| Strongly agree | 24 | 31 | 30 |
| Agree | 18 | 14 | 15 |
| Neither agree nor disagree | 2 | 1 | 1 |
| Disagree | 2 | 0 | 0 |
| Strongly disagree | 0 | 0 | 0 |

*Figure 5. Public stakeholders’ views about how advance research planning should inform decisions about research*

|  | Legally binding | Directive | Advisory |
| --- | --- | --- | --- |
| Strongly agree | 5 | 14 | 8 |
| Agree | 11 | 24 | 23 |
| Neither agree nor disagree | 7 | 4 | 6 |
| Disagree | 18 | 3 | 7 |
| Strongly disagree | 4 | 0 | 0 |

*Figure 6. Professional stakeholders’ views about how advance research planning should inform decisions about research*

|  | Legally binding | Directive | Advisory |
| --- | --- | --- | --- |
| Strongly agree | 5 | 14 | 7 |
| Agree | 11 | 23 | 23 |
| Neither agree nor disagree | 7 | 4 | 6 |
| Disagree | 17 | 3 | 7 |
| Strongly disagree | 4 | 0 | 0 |

*Figure 7. Public stakeholders’ views about what form advance research planning should take*

|  | Informal discussion | Formal process | Legal process |
| --- | --- | --- | --- |
| Strongly agree | 40 | 96 | 50 |
| Agree | 83 | 121 | 66 |
| Neither agree nor disagree | 73 | 37 | 76 |
| Disagree | 66 | 14 | 63 |
| Strongly disagree | 7 | 1 | 14 |

*Figure 8. Professional stakeholders’ views about what form advance research planning should take*

|  | Informal discussion | Formal process | Legal process |
| --- | --- | --- | --- |
| Strongly agree | 1 | 14 | 2 |
| Agree | 14 | 26 | 11 |
| Neither agree nor disagree | 14 | 4 | 16 |
| Disagree | 15 | 1 | 14 |
| Strongly disagree | 1 | 0 | 2 |

*Figure 9. Public stakeholders’ views about who should be involved in advance research planning*

|  | Person close | Dr/HCP from healthcare team | Dr/professional from research team |
| --- | --- | --- | --- |
| Strongly agree | 147 | 64 | 46 |
| Agree | 108 | 111 | 106 |
| Neither agree nor disagree | 11 | 69 | 82 |
| Disagree | 2 | 23 | 31 |
| Strongly disagree | 1 | 2 | 4 |

*Figure 10. Professional stakeholders’ views about who should be involved in advance research planning*

|  | Person close | Dr/HCP from healthcare team | DR/professional from research team |
| --- | --- | --- | --- |
| Strongly agree | 20 | 10 | 6 |
| Agree | 16 | 17 | 16 |
| Neither agree nor disagree | 8 | 12 | 14 |
| Disagree | 1 | 5 | 7 |
| Strongly disagree | 0 | 1 | 2 |
